# Metabolomic and Lipidomic Signatures of Depressive Symptoms in the REasons for Geographic And Racial Differences in Stroke (REGARDS) Study

**DOI:** 10.64898/2026.08.10.26360141

**Authors:** Chaitali Dagli, Nicole D. Armstrong, Pranali G. Patel, Eric Adjei Boakye, Zsuzsanna Ament, Amelia Demopoulos, Hemant K. Tiwari, Virginia J. Howard, W. Taylor Kimberly, Marguerite R. Irvin

## Abstract

**Background:** Depression is a highly prevalent condition with a substantial health burden and known links to metabolic dysfunction. We investigated plasma metabolites and lipids associated with depressive symptoms at baseline, including sex- and race-specific differences, and subsequently examined longitudinal associations of baseline metabolites and lipids with depressive symptoms over follow-up.

**Methods:** Baseline plasma samples were profiled for 162 metabolites and 195 lipid species using targeted mass spectrometry in the REasons for Geographic And Racial Differences in Stroke (REGARDS) stroke case-cohort. Depressive symptoms were assessed using the 4-item Center for Epidemiologic Studies Depression Scale (CES-D-4) at baseline and during follow-up. Cross-sectional associations were evaluated using weighted logistic regression, and longitudinal associations were assessed using weighted generalized estimating equations. Effect modification by sex and race was examined. Model 1 was adjusted for age, race, and sex (and time in longitudinal analyses). Model 2 was further adjusted for body mass index, smoking status, alcohol use, physical activity, perceived stress, and incident stroke. False discovery rate (FDR) correction was applied to account for multiple testing.

**Results:** Of 1,938 participants, 205 had depressive symptoms at baseline. At baseline, several metabolites (e.g., glycine, cyclic AMP) and lipid species were nominally associated with depressive symptoms, though none remained significant after FDR correction. In longitudinal analyses, stronger and more consistent associations emerged. Higher levels of anserine (OR=1.27, 95% CI: 1.16-1.39) and glucose (OR=1.36, 95% CI: 1.10-1.68) were associated with increased odds of depressive symptoms, with anserine remaining significant after FDR correction. Multiple lipid species, particularly phosphatidylethanolamines (PEs) and triglycerides (TGs), were significantly associated with depressive symptoms after FDR correction in longitudinal models. Significant sex interactions were observed for several lipid species, with stronger positive associations in females and attenuated or inverse associations in males.

**Conclusions:** In this stroke-enriched case-cohort, PEs and TGs were associated with depressive symptoms over time. Sex-specific lipid differences highlight biological heterogeneity. These results identify amino acid, phospholipid, and triacylglycerol pathways in depressive symptoms, supporting further mechanistic and biomarker investigations.

## Introduction

Depression is a prevalent and debilitating mental health condition that affects over 280 million people globally,^1^ contributing to a substantial burden on healthcare systems and society. It is characterized by persistent sadness, loss of interest or pleasure, and various physical and cognitive impairments.^2^ Depression is also associated with increased risk for chronic conditions such as cardiovascular disease, diabetes, and stroke,^3–5^ further emphasizing its impact on overall health. Females are about 50% more likely to experience depression than males,^6^ potentially due to a combination of biological, hormonal, and psychosocial factors.^7,8^ Additionally, depression is more prevalent among Black populations compared to other racial groups;^9^ however, these populations remain underrepresented in depression research, limiting insights into potential racial differences in biological pathways linked to depression. The growing need to understand the biological underpinnings of depression has sparked interest in the metabolomics and lipidomics fields to identify circulating plasma biomarkers and metabolic pathways associated with depression and depressive symptoms.

Emerging evidence suggests that metabolic dysregulation plays a critical role in the pathophysiology of depression.^10,11^ Several studies have also found links between depression and changes in plasma lipids such as fatty acids, phosphatidylcholines (PC), sphingolipids, cholesterol esters, phosphatidylethanolamines (PE), phosphatidylinositols, diacylglycerols, and triacylglycerols (TGs) across various populations.^11–13^ However, many of these studies were limited by small sample sizes and low coverage of the blood lipidome, leaving gaps in understanding the role of lipids in depressive symptoms. Furthermore, studies have also consistently reported alterations in metabolic pathways, including energy metabolism,^14^ lipid profiles,^15^ and amino acid processing,^16^ among individuals with depressive symptoms. For instance, in an untargeted mass spectrometry-based metabolomics study that measured over 800 compounds across large cohorts (n = 13,596) from various studies, researchers identified 8 metabolites linked to depression and depressive symptoms. After adjusting for lifestyle factors and the use of cardiovascular and antidepressant medications, higher levels of retinol (vitamin A), 1-palmitoyl-2-palmitoleoyl-GPC (16:0/16:1) (lecithin), and mannitol/sorbitol, as well as lower levels of hippurate, 4-hydroxycoumarin, 2-aminooctanoate (alpha-aminocaprylic acid), 10-undecenoate (11:1n1) (undecylenic acid), and 1-linoleoyl-GPA (18:2) (lysophosphatidic acid; LPA 18:2) were observed in individuals with depression.^16^ In the Finnish Depression and Metabolic Syndrome in Adults (FDMSA) cohort, certain metabolites, such as glucose, glycoprotein acetylation, creatinine, and TGs in very large high-density lipoproteins (XL-HDL-TG), were found to be associated with depressive symptoms.^17^ However, none of these associations were significant after adjusting for multiple comparisons. Network analysis found strong correlations between metabolites, but no direct influence on depressive symptoms, highlighting a complex relationship without clear causality.^17^ Furthermore, this research has mostly been based in European populations, therefore expansion to other groups remains necessary.

Despite advances in metabolomics and lipidomics research, inconsistencies remain in the literature regarding the association between metabolomic and lipid markers with depressive symptoms. Understanding these markers could enhance our ability to identify at-risk individuals. Therefore, in this study, we aimed to identify plasma metabolites and lipids associated with depressive symptoms in a case-cohort study of Black and White participants from the REasons for Geographic And Racial Differences in Stroke (REGARDS) cohort. We assessed associations at baseline and over follow-up to capture temporal patterns, with additional evaluation of sex-and race-specific differences at baseline. Our goal was to determine whether specific metabolites and lipids reflect underlying biological pathways relevant to depressive symptoms.

## Methods

### Study population

The REGARDS study is a national, population-based cohort designed to investigate racial and regional disparities in stroke incidence in the US. Between 2003 and 2007, REGARDS enrolled 30,239 non-Hispanic Black and White adults aged 45 years or older. Participants were recruited from across the continental US, with intentional oversampling of Black individuals and residents of the southeastern region, commonly referred to as the “stroke belt,” which includes Louisiana, Arkansas, Mississippi, Alabama, Tennessee, Georgia, North Carolina, and South Carolina, as well as the “stroke buckle” along the coastal plains of North Carolina, South Carolina, and Georgia. Detailed descriptions of the study design and recruitment procedures have been published previously.^18^ At baseline, participants completed a structured computer-assisted telephone interview (CATI) to collect demographic, clinical, and lifestyle information after providing verbal informed consent. This was followed by an in-home examination during which trained personnel obtained physical measurements and written informed consent and collected fasting blood samples via venipuncture. Blood samples were processed using standardized protocols and stored at −80 °C in a central laboratory for future analyses.^19^ Participants are followed by CATI every six months (through 03/2024 for these analyses) for suspected stroke events and general health information included depressive symptoms.

For this study, we used the REGARDS stroke case-cohort, a nested design including all adjudicated incident ischemic stroke cases accrued through April 1, 2019, along with a stratified random subcohort selected from the full REGARDS population to ensure representation across age, sex, and race groups.^20,21^ Participants with cognitive impairment defined as a Six-item Screener (SIS)^22^ score of less than or equal to 4 or a history of stroke at baseline were excluded from all analyses. For longitudinal analyses, participants without follow-up data on depressive symptoms were additionally excluded **(Supplementary Figure F1)**.

### Exposures

Metabolomic profiling was conducted using baseline plasma samples collected as part of an ancillary study within the REGARDS stroke case-cohort.^21^ EDTA plasma samples underwent targeted metabolomic profiling of aqueous compounds using dual Agilent 1290 Infinity II high-performance liquid chromatography systems coupled with a 6495 triple quadrupole tandem mass spectrometer (Agilent Technologies, Santa Clara, CA), as previously described.^20,23^ A total of 162 metabolites were quantified using this targeted platform.

Lipidomic profiling was performed using the same chromatographic and mass spectrometric instrumentation. Lipids were extracted from 10 μL plasma aliquots using 190 μL of isopropanol containing 1,2-didodecanoyl-sn-glycero-3-phosphocholine as an internal standard (Avanti Polar Lipids, Alabaster, AL).^24^ Following centrifugation at 9000 × g for 10 minutes at 4°C, supernatants were injected directly onto a Kinetex 100 × 2.1 mm (2.6 μm) column (Phenomenex, Torrance, CA). The mass spectrometer was configured to quantify 195 plasma lipid species, including both polar and nonpolar lipids, which were annotated by lipid class, total carbon number, and degree of unsaturation. Chromatographic separation was achieved using a binary solvent system consisting of mobile phase A (0.1% formic acid and 10 mM ammonium formate in water) and mobile phase B (0.1% formic acid in acetonitrile:isopropanol, 10:90 v/v).

The gradient increased from 30% to 99% mobile phase B over 17 minutes at a flow rate of 0.35 mL/min. Due to the non-normal distribution, all values were standardized before statistical analyses.

### Outcome

Depressive symptoms were assessed using the CES-D-4 scale. This abbreviated instrument was derived from the original 20-item CES-D and demonstrates reliability and validity comparable to the full-scale measure, with acceptable correlations with both the 20-item and 8-item versions.^25,26^ The CES-D-4 was administered via CATI at baseline and was at approximately 2-year intervals during follow-up. Each item was scored on a 4-point Likert scale ranging from 0 (“rarely or none of the time”) to 3 (“most or all of the time”), yielding a total score ranging from 0 to 12. Scores greater than or equal to 4 were used in analyses to define elevated depressive symptoms, reflecting clinically meaningful depressive symptomatology.^25^ Baseline CES-D-4 scores were used for cross-sectional analyses, while repeated CES-D-4 assessments obtained during follow-up were used for longitudinal analyses. Participants were required to have at least one follow-up assessment of depressive symptoms to be included in longitudinal analyses.

### Covariates

Demographic, behavioral, and clinical characteristics were included as covariates in all analyses. Age and body mass index (BMI) were modeled as continuous variables. Race was self-reported as Black or White, and sex was self-reported as female or male. Alcohol use was classified as current, past, or never. Smoking status was categorized as current smoker and non-smoker. Physical activity was assessed using the Life’s Simple 7 physical activity metric and categorized according to American Heart Association criteria (poor, intermediate, or ideal).^27^ Perceived stress was assessed using Cohen’s 4-item Perceived Stress Scale^28^ and categorized into tertiles (T1 ≤ 1, T2 = 2–4, T3 ≥ 5). Incident stroke cases were coded as yes or no. Antidepressant use was categorized as yes or no and was determined through an in-home medication bottle review.^29^

### Statistical Analyses

Weighted models were used to account for the stratified sampling design and to provide population-representative estimates of associations.

Descriptive analyses were conducted to summarize baseline demographic and clinical characteristics by baseline depressive symptom status. Continuous variables, including age and BMI, were summarized as means with standard deviations and compared using t-tests.

Categorical variables, including race, sex, alcohol use, smoking status, physical activity, perceived stress, and incident stroke cases, were summarized as counts and percentages and compared using chi-square tests.

Associations between each baseline metabolite or lipid and baseline depressive symptoms were evaluated using weighted logistic regression. For each exposure, odds ratios (ORs) and corresponding 95% confidence intervals (CIs) were estimated. Model 1 was adjusted for age, race, and sex, while Model 2 included additional adjustments for BMI, smoking status, alcohol use, physical activity, perceived stress, and incident stroke.

Associations between each baseline metabolite or lipid and repeated measures of depressive symptoms were assessed using weighted generalized estimating equations (GEE) with a logit link and an exchangeable working correlation structure to account for within-participant correlation across repeated measures. Study contacts for depressive symptoms, which occurred approximately every two years, were included as the time variable. For each metabolite and lipid, regression coefficients and robust standard errors were obtained from the GEE models and exponentiated to derive ORs with 95% CIs to facilitate interpretability and comparability with cross-sectional results. Model 1 adjusted for age, race, sex, and time and Model 2 additionally adjusted for BMI, smoking, alcohol use, physical activity, perceived stress, and incident stroke.

Potential effect modification by race and sex was evaluated for all metabolites and lipids using multiplicative interaction terms in weighted logistic regression models for baseline depressive symptoms in fully adjusted models.

To account for multiple comparisons, false discovery rate (FDR) correction was applied separately for metabolite and lipid analyses, with associations considered statistically significant at an FDR-adjusted *p* < 0.05. For interaction, a significance threshold of FDR < 0.10 was applied given the lower statistical power typically observed for interaction tests.^30^ All statistical tests were two-sided. All analyses were conducted using SAS v.9.4 (SAS Institute Inc., Cary, NC)

### Sensitivity Analyses

To assess the influence of antidepressant use on the observed associations, sensitivity analyses were conducted for the top five metabolites and top five lipids identified in the primary analyses. Weighted logistic regression and weighted GEE models were fitted with baseline antidepressant use included as an additional covariate in both Model 1 and Model 2.

To assess the robustness of primary associations, sensitivity analyses were conducted in the random subcohort, excluding participants with incident stroke. Associations between each baseline metabolite or lipid and baseline depressive symptoms were evaluated using the same weighted logistic regression approach and covariate adjustments as the main analyses. Analyses were restricted to baseline observations due to the limited sample size and number of repeated measures.

Exploratory pathway enrichment analyses were performed to aid the biological interpretation of the findings using MetaboAnalyst 6.0.^31^ Over-representation analyses were used to identify pathways enriched among the top 50 metabolites and lipids from each model. For metabolites, the Small Molecule Pathway Database (SMPDB) library containing 99 human metabolic pathways was used, whereas lipid enrichment analyses utilized the Lipidomics library (817 entries), integrating PathBank, Reactome, WikiPathways, and KEGG.

## Results

### Characteristics of Participants

**Table 1** presents the baseline demographic and clinical characteristics of participants in the REGARDS stroke case-cohort stratified by baseline depressive symptom status. Of the 1,938 participants, 205 had depressive symptoms at baseline. Among the 1,424 participants included in the longitudinal analyses, the median follow-up was 6 years. Participants with depressive symptoms were younger on average than those without depressive symptoms (mean age 64 vs. 68 years, p < 0.001). A higher proportion of participants with depressive symptoms were female (69.8% vs. 47.4%, p < 0.001) and Black (49.8% vs. 41.0%, p = 0.017). Participants with depressive symptoms were more likely to be current smokers (28.3% vs. 13.8%, p < 0.001) and reported lower physical activity (p = 0.0014). Participants with depressive symptoms had slightly higher BMI (mean 30 kg/m² vs. 29 kg/m², p = 0.011) and also reported higher perceived stress, with 76.1% in the highest tertile (T3) compared with 24.4% of those without depressive symptoms (p < 0.001).

**Table 1:** Demographic characteristics of REGARDS case cohort participants (N=1,938)

|  | Depression Scale |  | p-value |
| --- | --- | --- | --- |
|  | CES-D-4<br>≥ 4<br>(n=205) | CES-D-4<br>≤3<br>(n=1733) |  |
|  | N (%) | N (%) |  |
| Age (Mean, SD) | 64 (11.0) | 68 (10.0) | <0.0001 |
| Race |  |  | 0.0168 |
| Black | 102 (49.8) | 711 (41.0) |  |
| White | 103 (50.2) | 1022 (59.0) |  |
| Gender |  |  | <0.0001 |
| Female | 143 (69.8) | 822 (47.4) |  |
| Male | 62 (30.2) | 911 (52.6) |  |
| Alcohol use |  |  | 0.3015 |
| Current | 91 (44.4) | 866 (49.9) |  |
| Never | 72 (35.1) | 530 (30.7) |  |
| Past | 42 (20.5) | 337 (19.5) |  |
| Current smoker |  |  | <0.0001 |
| No | 147 (71.7) | 1487 (85.8) |  |
| Yes | 58 (28.3) | 239 (13.8) |  |
| Physical activity |  |  | 0.0014 |
| Poor | 88 (42.9) | 563 (32.5) |  |
| Intermediate | 76 (37.1) | 615 (35.5) |  |
| Ideal | 40 (19.5) | 524 (30.2) |  |
| Perceived stress |  |  | <0.0001 |
| T1 | 11 (5.4) | 686 (39.6) |  |
| T2 | 38 (18.5) | 624 (36.0) |  |
| T3 | 156 (76.1) | 423 (24.4) |  |
| Incident stroke cases |  |  | 0.6700 |
| No | 95 (46.3) | 776 (44.8) |  |
| Yes | 110 (53.7) | 957 (55.2) |  |
| BMI (Mean, SD) | 30 (7.0) | 29 (6.0) | 0.0105 |

Perceived stress tertiles: T1 ≤1; T2 = 2-4; T3 ≥5

### Metabolites and Depressive Symptoms

The top five associations between metabolites and depressive symptoms at baseline and over follow-up are presented in **Table 2**, with full results for all 162 metabolites provided in **Supplementary Tables S1 and S2**.

**Table 2:** Cross sectional and longitudinal association between metabolites with depressive symptoms.

| Model 1 |  |  |  | Model 2 |  |  |  |
| --- | --- | --- | --- | --- | --- | --- | --- |
| Cross sectional |  |  |  |  |  |  |  |
| Exposure | OR (95% CI) | p-value | FDR adjusted p-value | Exposure | OR (95% CI) | p-value | FDR adjusted p-value |
| Glycine | 0.70 (0.56, 0.88) | 2.43E-03 | 0.0946 | Glycine | 0.62 (0.46, 0.83) | 1.63E-03 | 0.1323 |
| Cyclic AMP | 1.46 (1.15, 1.85) | 1.99E-03 | 0.0946 | Cyclic AMP | 1.49 (1.16, 1.90) | 1.63E-03 | 0.1323 |
| Gluconic acid | 1.43 (1.16, 1.77) | 9.25E-04 | 0.0946 | Serine | 0.68 (0.53, 0.88) | 2.96E-03 | 0.1597 |
| Glycochenodeoxycholic acid | 1.31 (1.10, 1.57) | 2.92E-03 | 0.0946 | Anserine | 1.45 (1.12, 1.87) | 4.05E-03 | 0.1642 |
| Indole-3-propionic acid (IPA) | 0.13 (0.04, 0.47) | 1.99E-03 | 0.0946 | C26 carnitine | 0.67 (0.48, 0.93) | 1.78E-02 | 0.2089 |
| Longitudinal* |  |  |  |  |  |  |  |
| Gluconic acid | 1.39 (1.14, 1.70) | 1.22E-03 | 0.0985 | Anserine | 1.27 (1.16, 1.39) | 6.46E-07 | 0.0001 |
| Glucose | 1.42 (1.15, 1.75) | 1.11E-03 | 0.0985 | Glucose | 1.36 (1.10, 1.68) | 5.09E-03 | 0.4125 |
| Uridine | 0.73 (0.58, 0.91) | 5.75E-03 | 0.1707 | C16 carnitine | 1.27 (1.05, 1.55) | 1.44E-02 | 0.4675 |
| Leucine | 1.32 (1.08, 1.60) | 6.32E-03 | 0.1707 | Acetylglutamate | 1.17 (1.03, 1.32) | 1.27E-02 | 0.4675 |
| Anserine | 1.35 (1.09, 1.67) | 5.99E-03 | 0.1707 | Orotic acid | 1.36 (1.08, 1.72) | 1.04E-02 | 0.4675 |

*Top 5 metabolites*
*Model 1 adjusted for age, race, and sex*
*Model 2 further adjusted for BMI, perceived stress, smoking status, alcohol intake, physical activity, and incident stroke cases*
*\* Additionally adjusted for time*

In cross-sectional analyses, glycine and cyclic AMP showed consistent associations with depressive symptoms across models, with similar effect estimates after full adjustment. Glycine was inversely associated with depressive symptoms (Model 1: OR = 0.70; 95% CI = 0.56-0.88; Model 2: OR = 0.62; 95% CI = 0.46-0.83), whereas cyclic AMP was positively associated (Model 1: OR = 1.46; 95% CI = 1.15-1.85; Model 2: OR = 1.49; 95% CI = 1.16-1.90).

Additional metabolites were nominally associated with depressive symptoms in Model 2, including serine, anserine, and C26 carnitine. None of the cross-sectional associations remained statistically significant after FDR correction.

In longitudinal analyses, several metabolites demonstrated consistent or nominal associations with depressive symptoms across models, with modest differences after full adjustment. Glucose was positively associated with depressive symptoms in both models (Model 1: OR = 1.42; 95% CI = 1.15-1.75; Model 2: OR = 1.36; 95% CI = 1.10-1.68). Anserine was also positively associated (Model 1: OR = 1.35; 95% CI = 1.09-1.67; Model 2: OR = 1.27; 95% CI = 1.16-1.39), with a stronger and more precise estimate in the fully adjusted model. Additional metabolites were nominally associated with depressive symptoms in Model 2, including C16 carnitine, acetylglutamate, and orotic acid. After FDR correction, only anserine remained statistically significant in the fully adjusted longitudinal model.

### Lipids and Depressive Symptoms

The top five associations between lipids and depressive symptoms at baseline and over follow-up are presented in **Table 3**, with full results for all 195 lipid species provided in **Supplementary Tables S3 and S4**.

**Table 3:** Cross sectional and longitudinal association between lipids with depressive symptoms.

| Model 1 |  |  |  | Model 2 |  |  |  |
| --- | --- | --- | --- | --- | --- | --- | --- |
| Exposure | OR<br>(95% CI) | p-value | FDR<br>adjusted<br>p-value | Exposure | OR<br>(95% CI) | p-value | FDR<br>adjusted<br>p-value |
| **Cross sectional** |  |  |  |  |  |  |  |
| LPE(22:4) | 1.29 (1.09, 1.52) | 3.02E-03 | 0.0830 | LPC(22:5) | 1.38 (1.12, 1.70) | 2.73E-03 | 0.0833 |
| PC(32:1) | 1.29 (1.08, 1.53) | 3.83E-03 | 0.0830 | PC(32:1) | 1.33 (1.10, 1.60) | 2.79E-03 | 0.0833 |
| PE(34:1) | 1.35 (1.11, 1.64) | 3.02E-03 | 0.0830 | PI(38:3) | 1.43 (1.14, 1.78) | 1.71E-03 | 0.0833 |
| PE(36:4) | 1.38 (1.12, 1.70) | 2.56E-03 | 0.0830 | PI(38:4) | 1.47 (1.17, 1.85) | 1.06E-03 | 0.0833 |
| PE(38:4) | 1.42 (1.16, 1.73) | 5.89E-04 | 0.0830 | PS(38:6) | 1.36 (1.11, 1.67) | 2.99E-03 | 0.0833 |
| **Longitudinal\*** |  |  |  |  |  |  |  |
| PE(34:1) | 1.37 (1.18, 1.59) | 4.81E-05 | 0.0024 | TG(58:7) | 1.11 (1.06, 1.16) | 4.46E-06 | 0.0009 |
| PE(38:5) | 1.47 (1.22, 1.78) | 4.24E-05 | 0.0024 | PE(38:5) | 1.41 (1.17, 1.70) | 3.12E-04 | 0.0305 |
| TG(51:3) | 1.45 (1.23, 1.72) | 1.56E-05 | 0.0024 | PE(38:6) | 1.37 (1.14, 1.65) | 8.75E-04 | 0.0427 |
| PE(34:1) | 1.37 (1.18, 1.59) | 4.81E-05 | 0.0024 | TG(51:3) | 1.36 (1.14, 1.63) | 7.22E-04 | 0.0427 |
| TG(58:12) | 1.09 (1.04, 1.14) | 1.38E-04 | 0.0054 | PE(34:1) | 1.30 (1.11, 1.52) | 1.35E-03 | 0.0439 |

*Top 5 lipids*
*Model 1 adjusted for time, age, race, and sex*
*Model 2 further adjusted for BMI, perceived stress, smoking status, alcohol intake, physical activity, and incident stroke cases*
*\*Additionally adjusted for time*
*Each lipid peak represents a distinct LC-MS/MS signal; some signals share the same total carbon and double bond composition due to structural isomers, positional ambiguity, or analytical artifacts. All 195 peaks were analyzed individually for statistical testing and multiple comparison correction*

In cross-sectional analyses, the strongest associations were observed among glycerophospholipids [PE, PC phosphatidylinositols (PI), phosphatidylserines (PS), and lysophospholipids (lysophosphatidylethanolamine (LPE), lysophosphatidylcholine (LPC))]. In Model 1, several PE and LPE species were associated with increased odds of depressive symptoms, with representative ORs ranging from 1.29 to 1.42. In Model 2, similar associations were observed for PC, PI, and PS species, with ORs ranging from 1.33 to 1.47. None of the lipid cross-sectional associations remained statistically significant after FDR correction.

In longitudinal analyses, lipid associations with depressive symptoms were more pronounced than in cross-sectional analyses. In Model 1, both PE and TG species demonstrated consistent positive associations, with ORs ranging from 1.09 to 1.47 which remained in model 2 with similar ORs ranging from 1.11 to 1.41. After FDR correction, multiple lipid species remained statistically significant in both models, predominantly comprising PE species and TGs.

### Effect modification and Sex-specific Associations

Potential effect modifications by sex and race were evaluated in fully adjusted cross-sectional models for all metabolites and lipid species **(Supplementary Tables S5 and S8)**. Ten lipid species showed a statistically significant interaction effect with sex on depression, prompting sex-stratified analyses to further examine these differential associations **(Figure 1)**. No significant interactions were observed by race.

**Figure 1:**
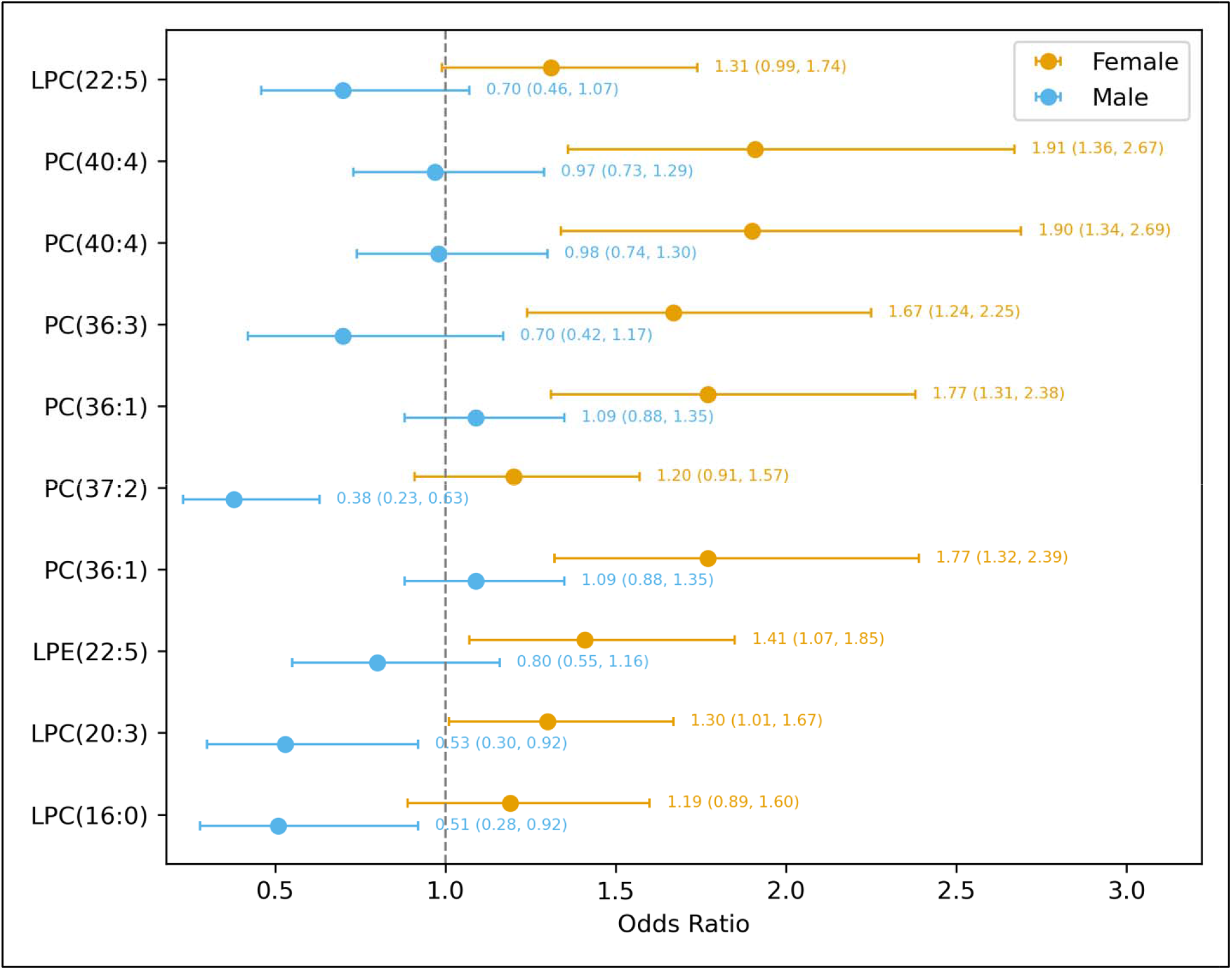
Lipid species subgroup analysis by sex. Stratified by sex Adjusted for age, race, BMI, perceived stress, alcohol intake, physical activity, and incident stroke FDR P for interaction: 0.1 Each lipid peak represents a distinct LC-MS/MS signal; some signals share the same total carbon and double bond composition due to structural isomers, positional ambiguity, or analytical artifact

In sex-stratified analyses **(Figure 1)**, several lipid species showed differing associations with depressive symptoms. Among lysophospholipids, LPC species demonstrated positive associations with depressive symptoms in females, for example, LPC(20:3) was associated with increased odds of depressive symptoms in females (OR = 1.30, 95% CI = 1.01-1.67), whereas an inverse association was observed in males (OR = 0.53, 95% CI = 0.30-0.92). Similarly, LPE species showed positive associations primarily in females, including LPE(22:5) (OR = 1.41, 95% CI = 1.07-1.85), with weaker associations in males. Multiple PC species also demonstrated stronger positive associations with depressive symptoms in females, including PC(36:1) (OR = 1.77, 95% CI = 1.32-2.39), PC(36:3) (OR = 1.67, 95% CI = 1.24-2.25), and PC(40:4) (OR = 1.90, 95% CI = 1.34-2.69), whereas corresponding associations in males were attenuated or inverse, such as PC(37:2) (OR = 0.38, 95% CI = 0.23-0.63). Overall, these findings indicate that associations between lipid species and depressive symptoms were more pronounced in females, while corresponding associations in males were weaker or in the opposite direction.

### Sensitivity Analyses

Additional adjustment for baseline antidepressant use had little impact on metabolite associations, all of which remained statistically significant. Among lipids, four of the top five associations identified in the cross-sectional Model 1 analyses were attenuated and no longer statistically significant after adjustment for antidepressant use. In contrast, all lipid associations identified in the fully adjusted cross-sectional Model 2 and longitudinal analyses remained statistically significant. Full results are presented in **Supplementary Tables S9 and S12**.

In the random subcohort, results remained consistent with the primary cross-sectional findings for both metabolites and lipid species; full results are presented in **Supplementary Tables S13 and S14**.

### Enrichment Analyses

Pathway enrichment analyses of the top 50 metabolites associated with depressive symptoms in the cross-sectional analyses primarily identified amino acid–related pathways, including glycine and serine metabolism and arginine and proline metabolism, along with nicotinate/nicotinamide metabolism and phosphatidylcholine biosynthesis in fully adjusted models. Longitudinal analyses further highlighted enrichment of energy-related pathways, including the citric acid cycle and mitochondrial electron transport. In contrast, lipid enrichment analyses did not identify statistically significant pathways after multiple testing correction, likely due to minimal overlap among the top lipid species and the predominance of non-specific drug-related pathway annotations. Full enrichment results are presented in the **Supplementary Results and Supplementary Figures F2 to F9**.

## Discussion

In this large, population-based sample of adults, we identified several plasma metabolites and lipid species associated with depressive symptoms at baseline and over follow-up. Baseline associations were modest and did not persist after correction for multiple testing; however, longitudinal analyses revealed stronger lipid associations, particularly among PEs and TGs, several of which remained significant after multiple testing correction. Among non-lipid metabolites, amino acid–related and energy-related compounds demonstrated consistent patterns across models, with anserine showing strong and consistent associations in longitudinal analyses. We also observed sex-specific lipid associations at baseline, with several PC and lysophospholipid species demonstrating differential associations in females and males. Pathway enrichment analyses of metabolites highlighted amino acid and energy metabolism pathways as recurrent themes, whereas lipid enrichment analyses did not identify statistically significant pathways.

### Metabolite Signatures of Depressive Symptoms in the Context of Energy Metabolism and Dietary Influences

In our longitudinal analyses, higher anserine levels were associated with increased odds of depressive symptoms. Supporting this finding, a case-control study found higher serum levels of carnosine, a related histidine-containing dipeptide (HCD), among individuals with major depressive disorder compared with controls (OR = 1.90, 95% CI = 1.22-2.94).^32^ Given that anserine is a methylated derivative of carnosine, these dipeptides are closely linked within HCD metabolism and may reflect shared metabolic and dietary influences.^33^ In contrast, a meta-analysis of randomized controlled trials conducted across diverse populations found that HCD supplementation (20 studies), including anserine, carnosine, L-carnosine, or β-alanine, demonstrated significant reductions in depression scores compared with placebo (mean difference = −0.79; 95% CI = −1.24, −0.35).^34^ Mechanistically, HCDs are thought to exert neuroprotective effects through antioxidant activity, buffering reactive oxygen species, and modulating glutamatergic and monoaminergic neurotransmission.^33,35^ The discrepancy between supplementation trials and observational findings may therefore reflect differences between exogenous and endogenous HCD levels, with higher baseline anserine potentially indicating dysregulated amino acid metabolism, altered renal clearance, or diet-related patterns linked to depressive symptomatology.

In parallel, higher glucose levels were associated with increased odds of depressive symptoms (Model 2: OR = 1.36, 95% CI = 1.10-1.68). Similar associations were reported in the FDMSA study, where a 1-SD higher glucose level was linked to higher Beck Depression Inventory scores (Model 1: β = 0.68, 95% CI: 0.18–1.17; Model 2: β = 0.49, 95% CI: −0.01– 1.0), although these associations attenuated after multiple testing adjustment.^17^ Likewise, a multi-cohort European study (N ≈ 12,586) observed a trend toward higher glucose levels being associated with depressive symptoms, suggesting that glucose may reflect subtle energy- and diet-related metabolic perturbations linked to mood regulation.^16^

Extending these findings, top metabolites in both cross sectional and longitudinal analyses spanning carbohydrate, nucleotide, and lipid metabolism, including gluconic acid, uridine, and long-chain acylcarnitines (C16 and C26) were associated with depressive symptoms. Together, these patterns are consistent with possible perturbations in oxidative stress, energy production, and mitochondrial function.^16,36,37^ These pathways are closely intertwined with both endogenous metabolic regulation and dietary exposures, such as intake of simple sugars, RNA-rich foods, and fat composition. This suggests that the observed associations may reflect a combination of altered biochemical processing and diet-related influences.

### Lipidomic Signatures of Depressive Symptoms in the Context of Immunometabolic and Dietary Factors

In contrast to metabolite findings, lipid species demonstrated more consistent longitudinal associations with depressive symptoms. Repeated-measures analyses identified 16 lipid species in minimally adjusted models and 6 in fully adjusted models that remained significant after correction. Associations were predominantly observed among PE species and TGs, with effect estimates ranging from approximately 1.1 to 1.5. These findings align with prior large-scale lipidomic analyses. In a pooled Dutch meta-analysis of approximately 15,400 individuals (5,283 cases), 21 lipid and lipoprotein-related markers including higher TGs, very-low-density lipoprotein cholesterol, apolipoprotein B, and glycoprotein acetyls, and lower high-density lipoprotein cholesterol were associated with depression, with modest effect sizes (pooled ORs per SD: ∼1.05-1.20).^38^ Similarly, in Bernhardsen *et al.* study (n = 777; 319 cases), several lipid species were nominally associated with depressive symptoms including TGs in very small very-low-density lipoproteins (VLDL) (XS-VLDL-TG) and XL-HDL-TG.^17^ These differences likely reflect variation in analytic platform (NMR lipoprotein profiling vs mass spectrometry– based lipidomics), lipid coverage, sample composition, and the incorporation of repeated-measures modeling in our study.

Biologically, PE and related phospholipid species are integral components of neuronal membranes and influence membrane fluidity, receptor localization, and synaptic signaling.^39,40^ In contrast, TG-related associations may reflect broader links between depressive symptoms and systemic metabolic dysregulation.^17,38,41^ Elevated TGs are a hallmark of insulin resistance and hepatic overproduction of VLDL, processes that are tightly coupled with chronic low-grade inflammation. This metabolic-inflammatory milieu is characterized by increased circulating cytokines (e.g., IL-6, TNF-α) and altered adipokine signaling, which can influence central nervous system function through multiple pathways, including disruption of the hypothalamic–pituitary–adrenal axis, altered monoamine metabolism, and impaired neuroplasticity.^17,42,43^ In addition, diet represents an important upstream determinant of these processes, as dietary patterns characterized by high intake of refined carbohydrates, saturated fats, and ultra-processed foods are associated with elevated TG levels, insulin resistance, and systemic inflammation.^38,41^ Within this framework, TG-rich lipoproteins may serve as peripheral markers of an underlying diet-sensitive immunometabolic state that contributes to depression risk rather than acting as direct causal agents. The persistence of these associations over time may reflect relatively stable metabolic or inflammatory states rather than transient fluctuations.

### Sex-Specific Lipid Signatures of Depressive Symptoms and Underlying Biological Differences

Notably, several PC and lysophospholipid species demonstrated stronger positive associations in females, with attenuated or inverse associations in males. A similar pattern was observed in the previously described analysis of large European cohorts, where effect sizes also tended to be larger in females, although formal evidence for sex-specific effect modification was not present.^16^ These findings align with clinical data suggesting sex-specific lipid alterations in depression. For example, a study of individuals with first-diagnosed, drug-naïve major depressive disorder reported sex-specific differences in erythrocyte fatty acid composition, with higher levels of n-6 polyunsaturated and saturated fatty acids and stronger associations with depressive symptom severity observed in females than males.^44^

Females generally exhibit higher subcutaneous adiposity and distinct lipoprotein characteristics, factors that may contribute to altered membrane lipid remodeling.^45,46^ Changes in the composition of phospholipids such as PCs and PEs can influence membrane fluidity and receptor organization, with downstream effects on neurotransmitter systems, including serotonin and dopamine, that are central to mood regulation.^47,48^ In addition, sex differences in immune function and hypothalamic–pituitary–adrenal axis activity may further shape these lipid– neurobiological interactions.^49^ Together, these mechanisms suggest that lipid perturbations associated with depressive symptoms may be differentially expressed by sex, although the magnitude and direction of these effects likely depend on underlying metabolic and population characteristics.

### Study Strength and Limitations

Several strengths of this study merit consideration. The analysis was conducted in a relatively large, community-based cohort of Black and White individuals, enhancing generalizability beyond predominantly European samples. The integration of both cross-sectional and longitudinal repeated-measures analyses allowed assessment of temporal consistency in metabolic associations. Comprehensive coverage of both intermediary metabolites and detailed lipid species enabled evaluation across multiple biochemical pathways. Rigorous control for multiple testing using FDR correction and the inclusion of sensitivity analyses reduced the likelihood of false-positive findings. Additionally, exploration of sex-specific associations provided insight into potential biological heterogeneity.

This study also has limitations. First, cross-sectional associations may be influenced by variability in baseline depressive symptom severity and transient physiological states.

Longitudinal models were also incorporated, but repeated metabolomic assessments were not available, restricting the ability to evaluate within-person metabolic change over time. Because 10.6% of participants had depressive symptoms at baseline and pre-enrollment depression history was not available, the observed associations may reflect current or prior depression-related biology rather than clear prospective risk markers. In addition, residual confounding cannot be excluded, as factors that may influence circulating metabolite and lipid concentrations, including dietary intake and renal function, were not included in the models, limiting causal interpretation of the observed associations. Second, not all metabolites and lipids among the top-ranked associations were represented in MetaboAnalyst for pathway enrichment analyses. This incomplete mapping may have limited the detection or interpretation of certain pathway-level signals; however, the principal enriched pathways were supported by multiple overlapping metabolites, suggesting that the overall pathway conclusions are unlikely to be driven by a single unmapped feature. Third, depressive symptoms were assessed using the CESD-4, which captures symptoms experienced during the prior week and does not distinguish between transient distress and chronic or recurrent depression, which may lead to misclassification of longer-term depressive status.

## Conclusion

These findings provide information on metabolic associations with depressive symptoms. Anserine was marginally associated with depression cross-sectionally and robustly with longitudinal measures after correction for multiple testing. Furthermore, longitudinal lipid associations were stronger compared to metabolites particularly for PE species and TGs. Notably, several lipid associations differed by sex, highlighting biological heterogeneity in metabolic correlates of depressive symptoms. The identification of amino acid, energy metabolism, and phospholipid pathways suggest candidate metabolite markers warranting evaluation in future studies.

## Supporting information

Supplemental Methods and Results

## Ethical Approval Declaration

The REGARDS study was approved by the Institutional Review Boards (IRBs) of the participating institutions, including the University of Alabama at Birmingham (UAB) and all collaborating institutions. The study was conducted in accordance with the ethical principles outlined in the Declaration of Helsinki.

## Data Availability

The datasets generated during and/or analysed during the current study are not publicly available due to participant privacy concerns. In order to abide by its obligations with NIH/NINDS and the UAB IRB, REGARDS facilitates data sharing through formal data use agreements. Any investigator is welcome to request the REGARDS data and documentation through this process. Requests for data access may be sent to the REGARDS study at.

## Author Contributions

C.D. conceptualized the study, performed the statistical analyses, interpreted the findings, and drafted the manuscript. N.D.A., P.G.P., E.A.B., and H.K.T. contributed to the interpretation of results and critically reviewed the manuscript. Z.A. and A.D. contributed to metabolomic and lipidomic data generation, processing, and interpretation, and critically reviewed the manuscript.

V.J.H. contributed to the REGARDS study design, interpretation of findings, and manuscript revision. W.T.K. contributed to the study conceptualization, data resources, interpretation of findings, and critical revision of the manuscript. M.R.I. supervised the study, contributed to study conceptualization and methodology, and critically reviewed the manuscript. All authors reviewed and approved the final version of the manuscript.

## Acknowledgment

The authors thank the other investigators, the staff, and the participants of the REGARDS study for their valuable contributions. A full list of participating REGARDS investigators and institutions can be found at: https://www.uab.edu/soph/regardsstudy/

