## Supplemental Methods and Results for "Metabolomic and Lipidomic Signatures of Depressive Symptoms in the REasons for Geographic And Racial Differences in Stroke (REGARDS) Study"

*Addressing multiple signals and structural ambiguity in lipidomics data*

Liquid chromatography-mass spectrometry (LC-MS/MS)-based lipidomics often encounters situations where multiple chromatographic signals correspond to the same nominal molecular formula, complicating accurate structural assignment. In our dataset, 195 chromatographic peaks were manually processed and integrated as distinct analytical signals. Among these, 142 compounds produced unique signals, while 53 displayed some level of analytical ambiguity.

Several factors contribute to the presence of similar or overlapping identifiers within the same dataset. For many lipid classes, particularly phosphatidylcholines (PC) and phosphatidylethanolamines (PE), positional isomers could not be determined, leading to signals representing the same fatty acid composition but with uncertain stereospecific numbering (sn), sn-1/sn-2 placement. Additionally, different structural variants can share the same molecular formula while differing in their fatty acid chain combinations. For example, a signal annotated as PC(38:4) may correspond to either PC(16:0_22:4) or PC(18:0_20:4). Beyond structural isomers, analytical artifacts may also produce multiple signals for the same compound, including chromatographic ionization variants (e.g., different adduct forms such as [M+H]^+^ or [M+NH_4_]^+^), matrix effects, that alter retention or ionization behavior, and column or instrument variations such as shifts in temperature, pressure, or flow rate.

In this dataset, although each of the 195 lipid peaks represents a unique chromatographic signal, analytical challenges and limitations result in cases where some distinct lipid signals share the same compositional sum of total carbon numbers and double bonds.

For traceability, the chromatographic peak identifiers assigned during manual integration are labelled as 1- 195, where each represents a distinct analytical signal regardless of the confidence of structural assignment. Next, species-level annotations are provided for compounds, where individual fatty acid chains within a lipid molecule were specified. Additionally, compositional sum identifiers are included to summarize the total carbon number and degree of unsaturation.

In statistical analyses, each lipid signal was tested, and 195 was used to correct for multiple testing.

**Results**

*Sensitivity Analyses*

Enrichment Analyses: Metabolites

Enrichment analyses of the top 50 metabolite associations with depressive symptoms also revealed largely consistent patterns across cross-sectional models **(Supplementary Figures F2–F5)**. For cross-sectional analyses, Model 1 highlighted several pathways, including phosphatidylcholine biosynthesis, methylhistidine metabolism, glycine and serine metabolism, methionine metabolism, histidine metabolism, ammonia recycling, glutathione metabolism, glutamate metabolism, beta-alanine metabolism, betaine metabolism, carnitine synthesis, arginine and proline metabolism, and phosphatidylethanolamine biosynthesis. Model 2 largely confirmed these findings, with additional emphasis on purine metabolism, nicotinate and nicotinamide metabolism, alanine metabolism, and histidine metabolism, reflecting modest shifts in pathway ranking between the minimally and fully adjusted models. Across both models, glycine/serine metabolism, phosphatidylcholine biosynthesis, and methylhistidine metabolism were consistently enriched, suggesting robust involvement of amino acid–related and phospholipid metabolic pathways in depressive symptomatology at baseline.

Longitudinal analyses using GEE models similarly identified overlapping pathways with the cross-sectional results. GEE Model 1 showed enrichment in glucose-alanine cycle, urea cycle, glycine and serine metabolism, alanine metabolism, beta-alanine metabolism, arginine and proline metabolism, pyrimidine metabolism, Warburg effect, methylhistidine metabolism, tryptophan metabolism, histidine metabolism, and phenylalanine and tyrosine metabolism. GEE Model 2 reinforced these findings with stronger signals in arginine and proline metabolism, urea cycle, pyrimidine metabolism, histidine metabolism, glycine and serine metabolism, alanine metabolism, glutathione metabolism, and Warburg effect, consistent with repeated measures capturing cumulative metabolic effects over time.

Overall, the enrichment results across cross-sectional and longitudinal analyses revealed convergent pathways in amino acid metabolism, nitrogen handling (urea/ammonia cycles), phospholipid biosynthesis, and energy metabolism, while fully adjusted and repeated-measures models highlighted additional intermediary metabolism and mitochondrial energy pathways.

Lipids

Enrichment analyses of the top 50 lipid associations with depressive symptoms also revealed largely consistent patterns across cross-sectional models **(Supplementary Figures F6–F9)**. In both Model 1 and Model 2, acetylcholine synthesis and phospholipid biosynthesis were identified; however, these pathways were supported by minimal overlap and did not remain statistically significant after correction for multiple comparisons. A broad set of drug-related pathways, including common non-steroidal anti-inflammatory drugs (NSAIDs), acetaminophen, and other pharmacologic action pathways, were also identified in both models, likely reflecting overlap between lipid species and pathway annotation databases rather than direct biological mechanisms. Overall, the enrichment profiles were highly similar between Model 1 and Model 2, with full covariate adjustment producing negligible changes in pathway ranking.

Enrichment analyses of the top lipid associations identified in longitudinal models using repeated measures showed a similar pattern. Both Model 1 and Model 2 again highlighted acetylcholine synthesis and phospholipid biosynthesis pathways, alongside numerous drug-related pathways; however, none of these pathways demonstrated statistical significance after multiple testing correction. No additional lipid-specific metabolic pathways were identified in longitudinal analyses, and the observed pathways were driven by limited overlap.
