## Supplemental Figures for "Metabolomic and Lipidomic Signatures of Depressive Symptoms in the REasons for Geographic And Racial Differences in Stroke (REGARDS) Study"

**Supplemental Figure 1:** Flow chart of REGARDS participants

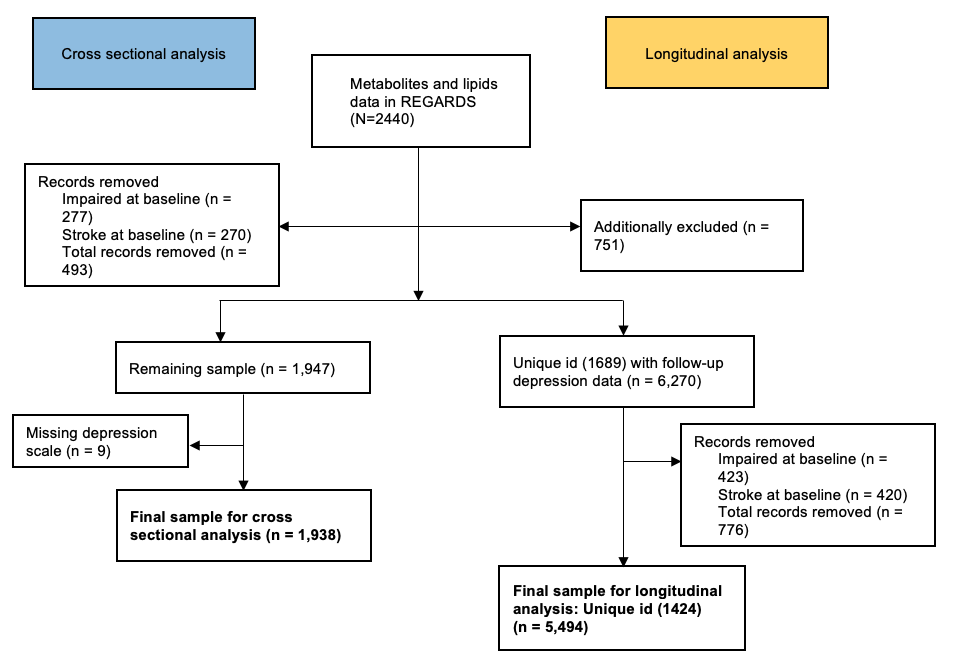

**Supplemental Figure 2:** Enrichment analyses of the top 50 metabolites associated with depressive symptoms in cross-sectional Model 1

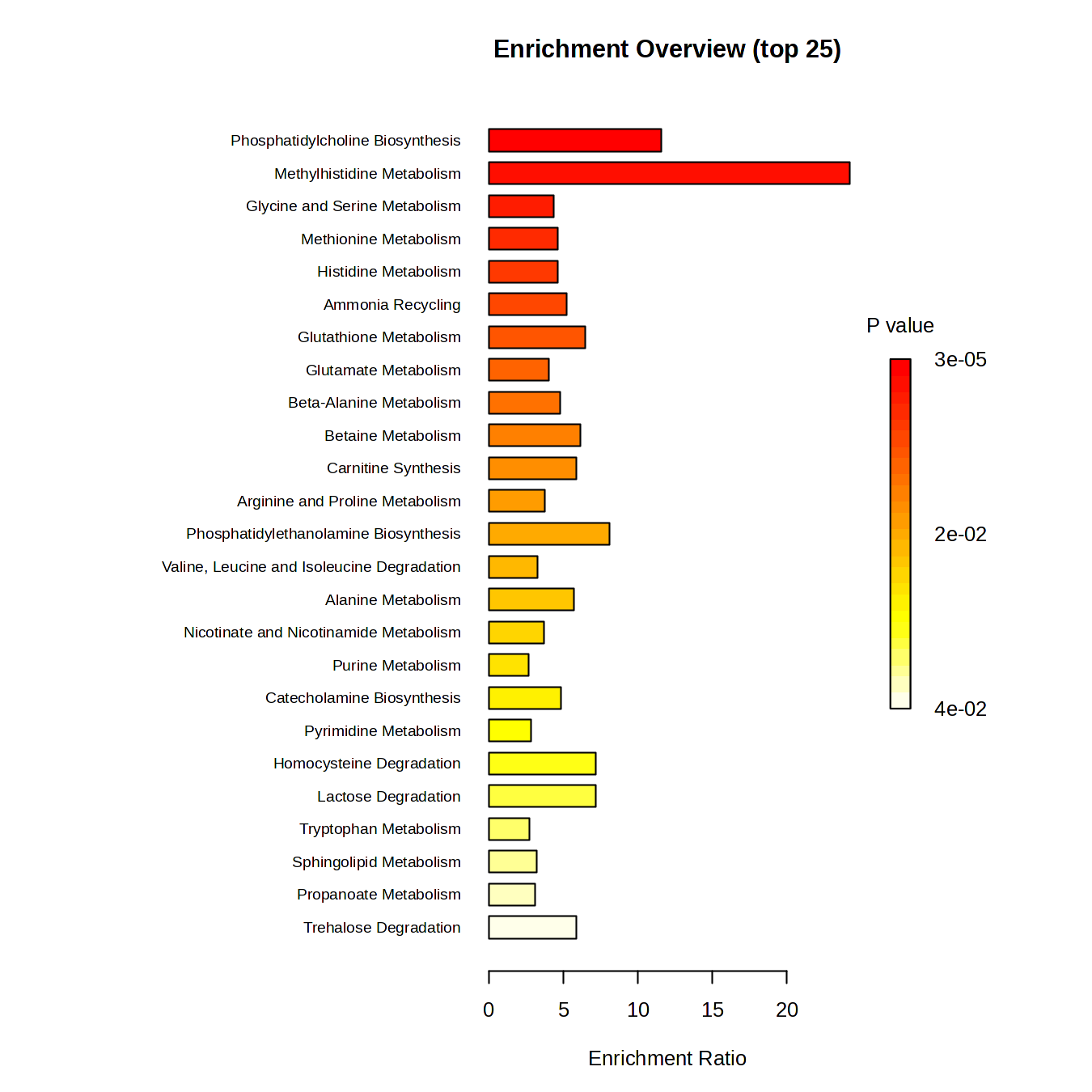

**Supplemental Figure 3:** Enrichment analyses of the top 50 metabolites associated with depressive symptoms in cross-sectional Model 2

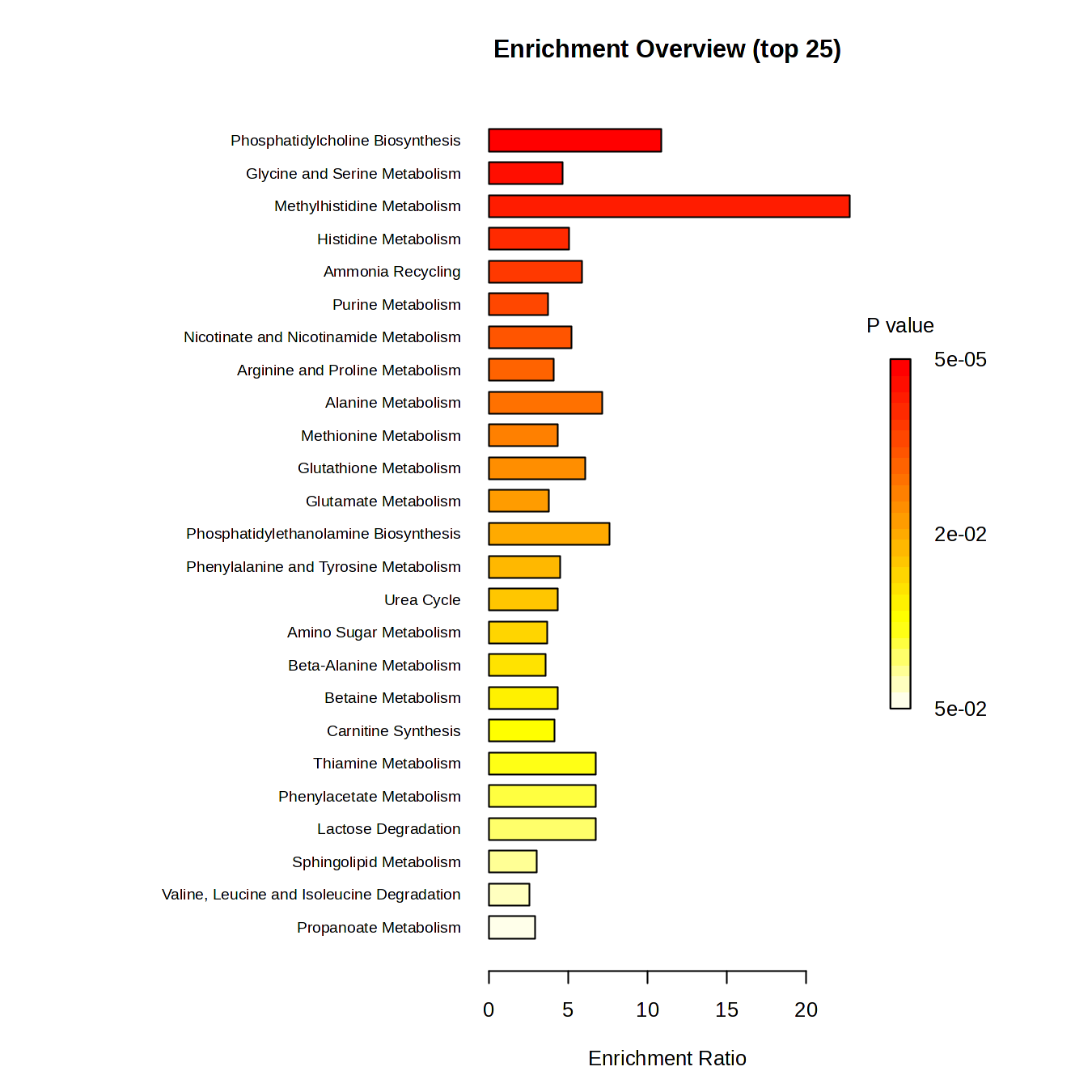

**Supplemental Figure 4:** Enrichment analyses of the top 50 metabolites associated with depressive symptoms in longitudinal Model 1

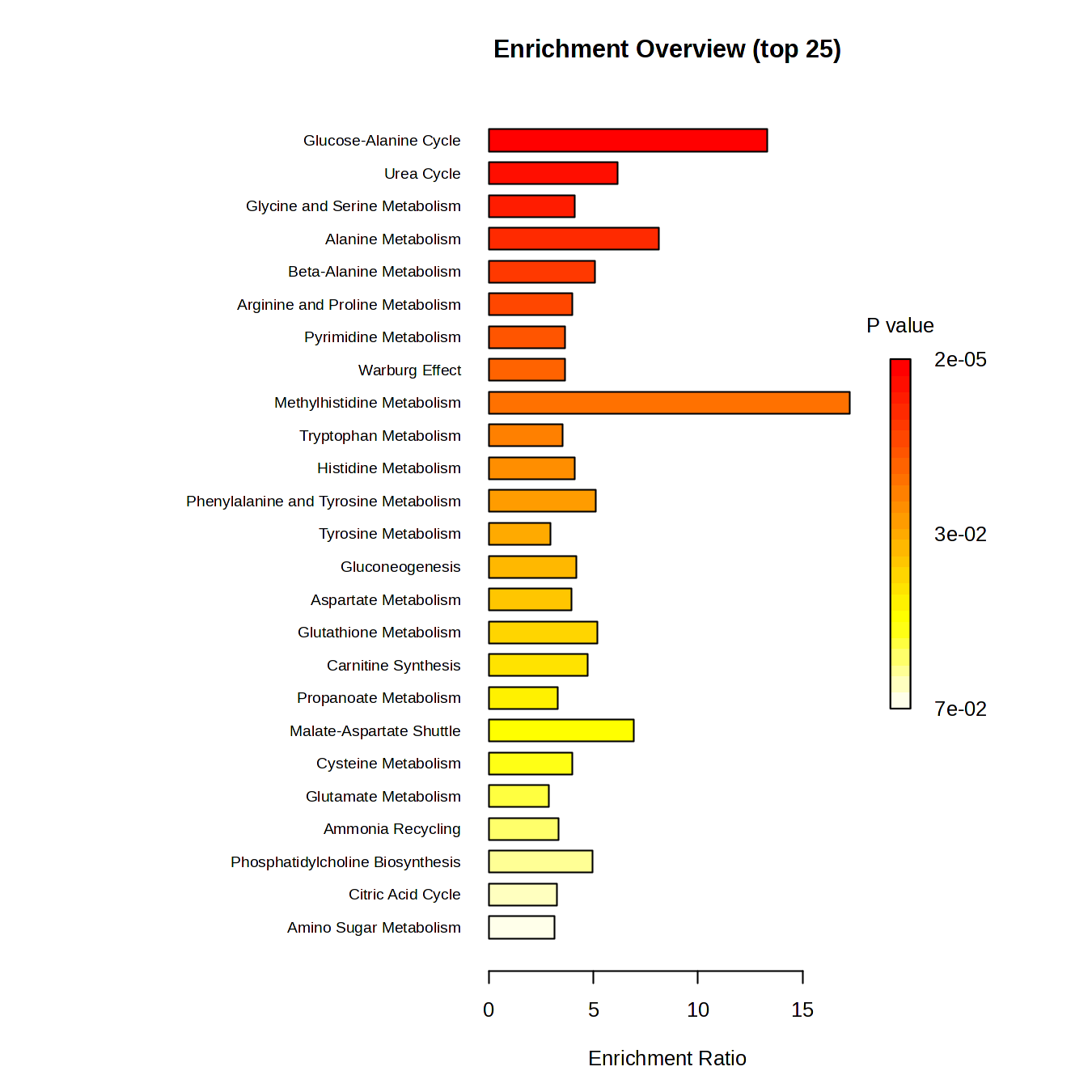

**Supplemental Figure 5:** Enrichment analyses of the top 50 metabolites associated with depressive symptoms in longitudinal Model 2

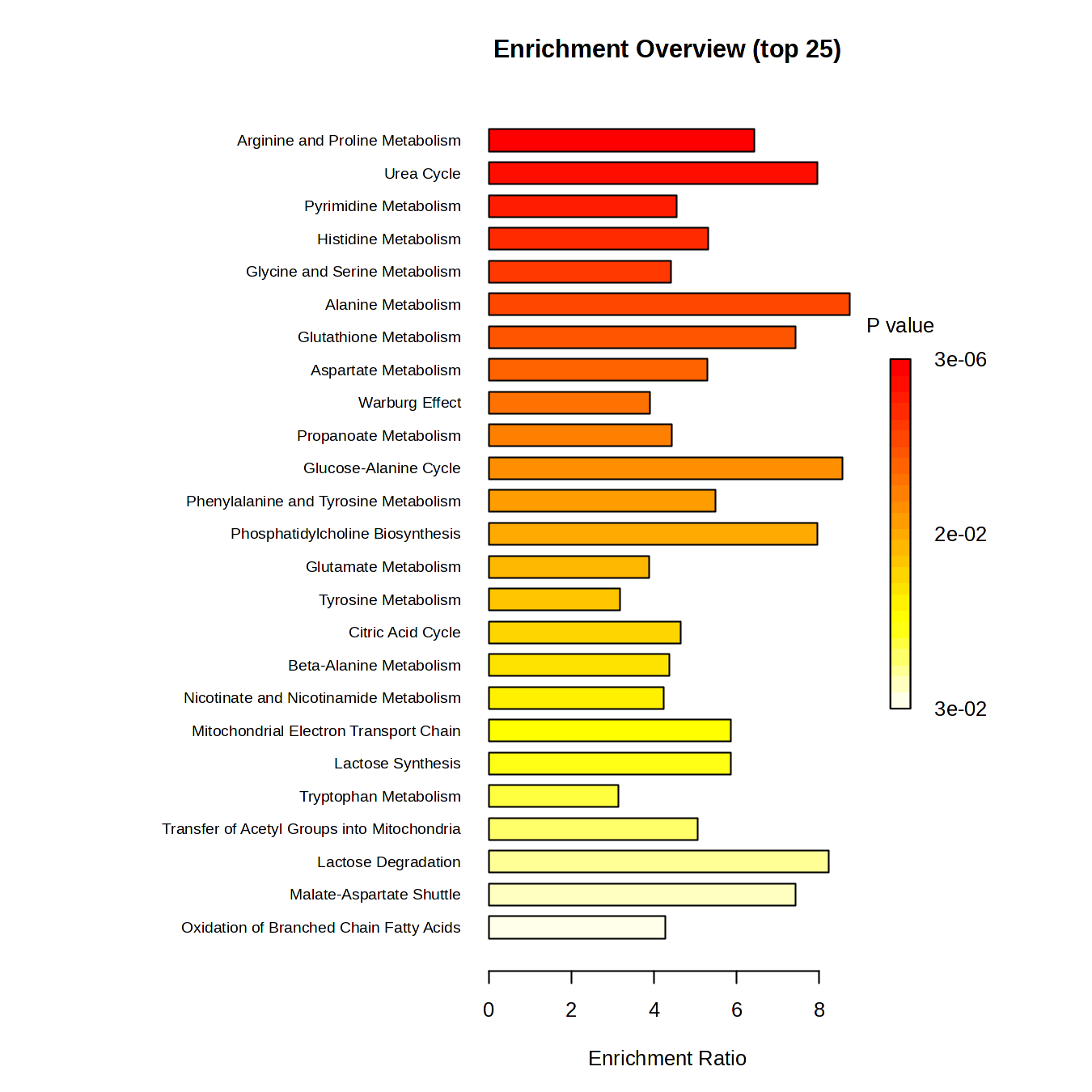

**Supplemental Figure 6:** Enrichment analyses of the top 50 lipids associated with depressive symptoms in cross-sectional Model 1

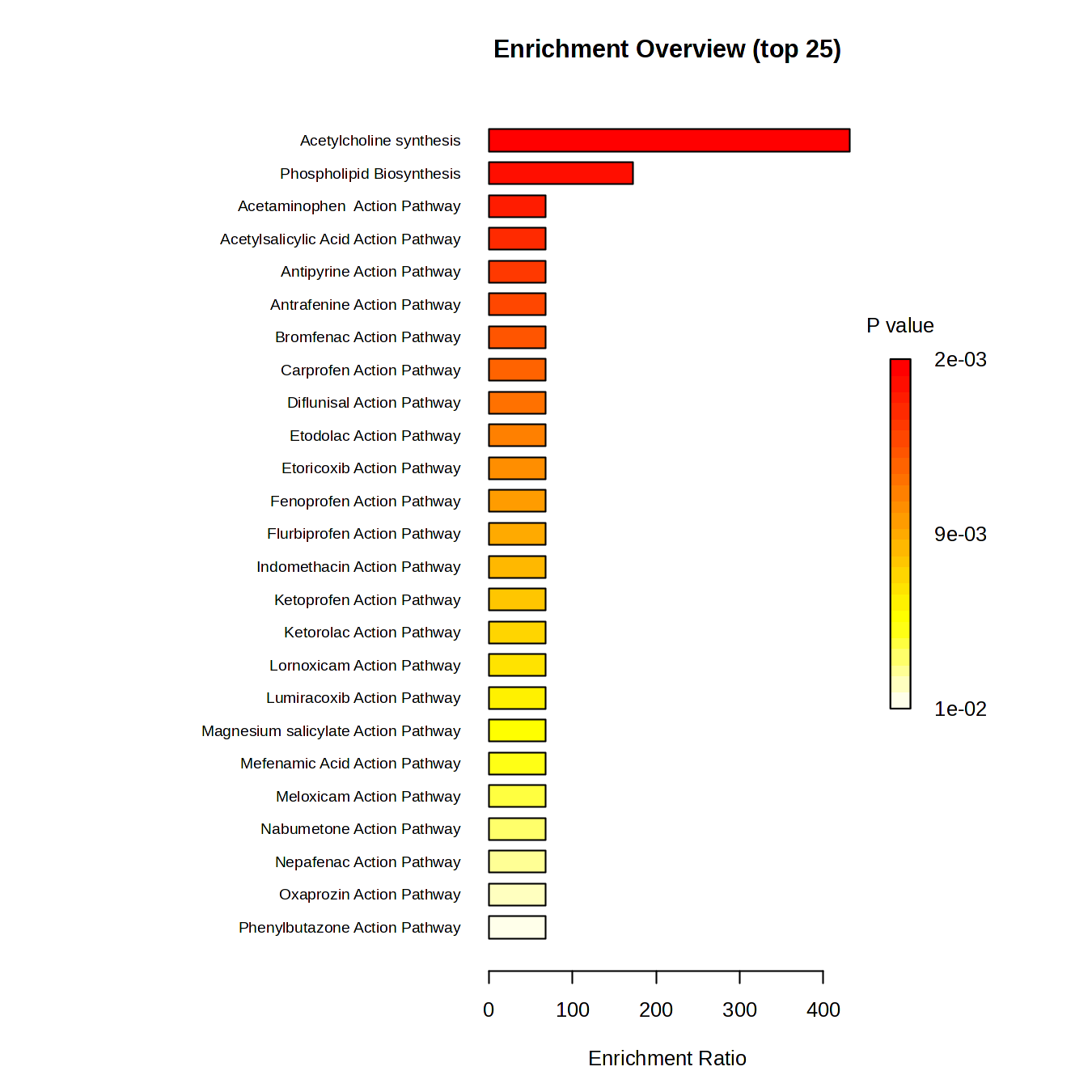

**Supplemental Figure 7:** Enrichment analyses of the top 50 lipids associated with depressive symptoms in cross-sectional Model 2

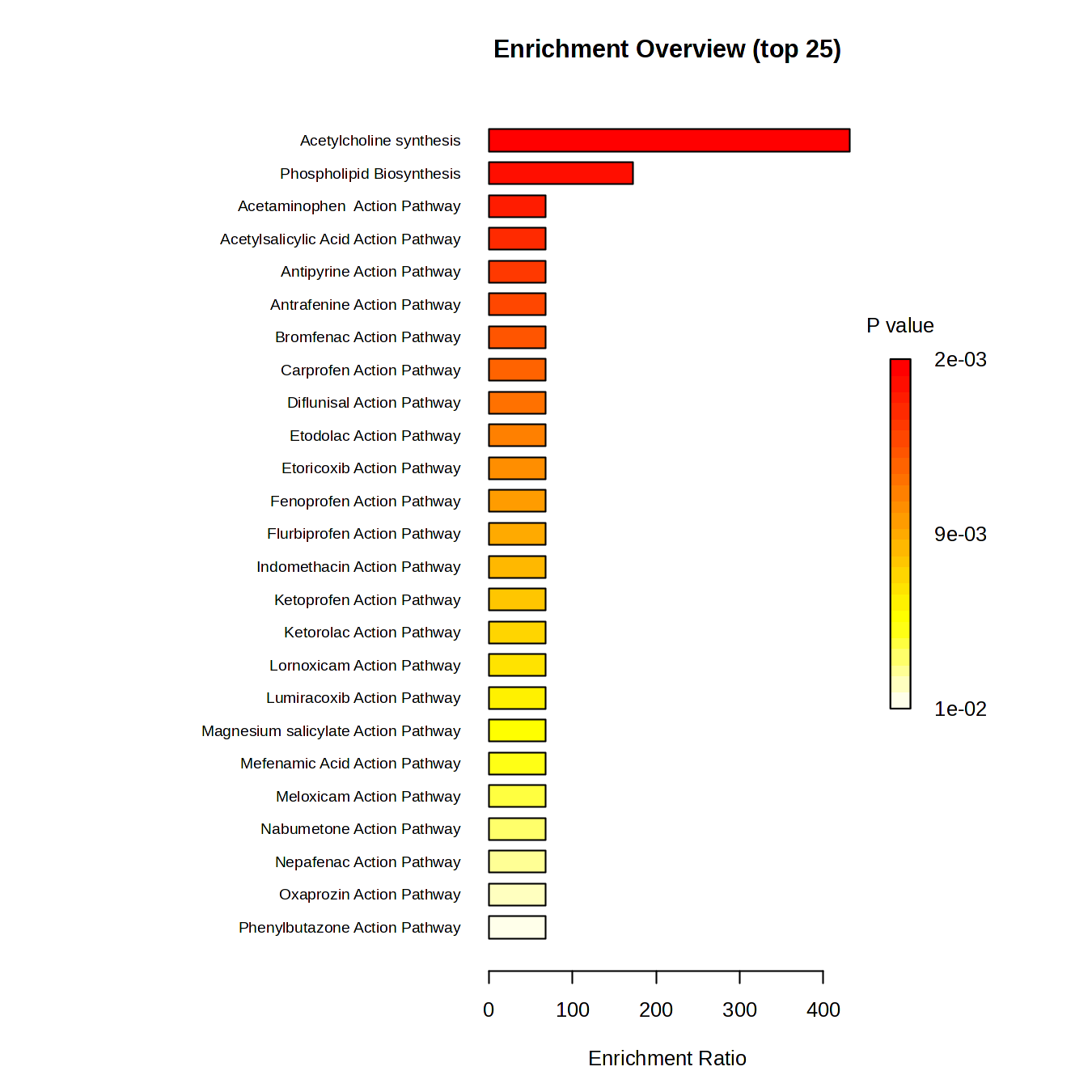

**Supplemental Figure 8:** Enrichment analyses of the top 50 lipids associated with depressive symptoms in longitudinal Model 1

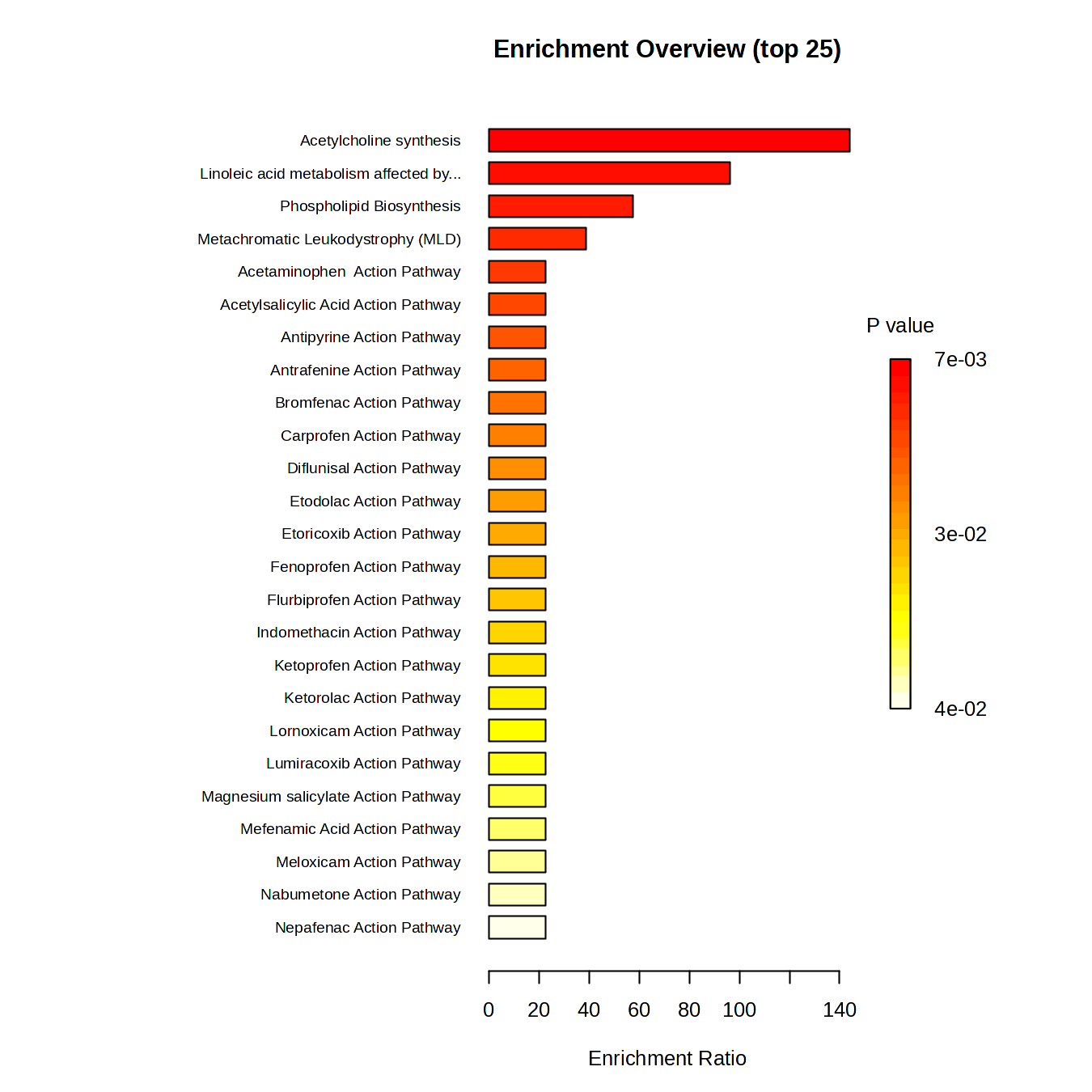

**Supplemental Figure 9:** Enrichment analyses of the top 50 lipids associated with depressive symptoms in longitudinal Model 2

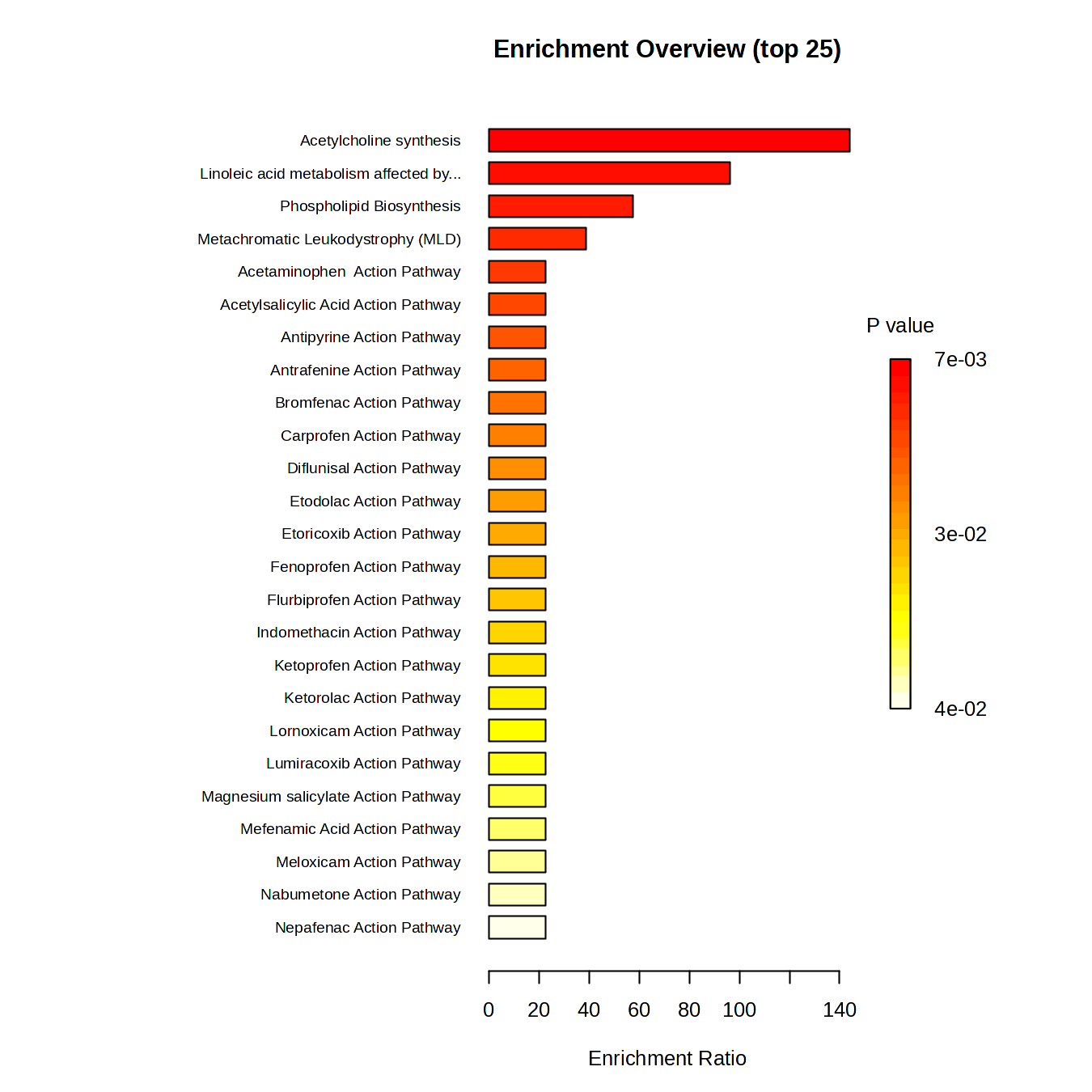
